# Sex differences in frailty and its impact on anticoagulation and hospitalization in older adults with atrial fibrillation

**DOI:** 10.1101/2025.05.15.25327673

**Authors:** Tu Nguyen, Tan Van Nguyen, Vinh Quang Nguyen, Huy Quoc Nguyen, Mark Woodward

**Affiliations:** The George Institute for Global Health, Australia; School of Public Health, The University of Sydney, Australia; Department of Geriatrics and Gerontology, University of Medicine and Pharmacy at Ho Chi Minh City, Vietnam; Department of Interventional Cardiology, Thong Nhat Hospital, Ho Chi Minh City, Vietnam; The George Institute for Global Health, School of Public Health, Imperial College London, United Kingdom

**Keywords:** Frailty, atrial fibrillation, anticoagulants, adverse outcomes, sex differences, gender differences

## Abstract

**Background:** Frailty is common in older patients, and it has been suggested to occur more frequently and with greater severity in women compared to men. In older people with atrial fibrillation (AF), frailty has been shown to influence the prescriptions of oral anticoagulants (OAC) and patient outcomes.

**Aim:** To examine the prevalence of frailty in older patients with AF and the association between frailty with OAC prescription and hospitalization, with a particular focus on variations by sex.

**Methods:** Adults aged ≥60 years with AF were recruited at the outpatient clinics of two major hospitals in Vietnam between December 2022 and May 2024. Frailty was defined by the 9-item Clinical Frailty Scale (CFS) with a cut-point of 5. Logistic regression models were applied to examine the association between CFS score with OAC prescription and cardiovascular disease (CVD) related hospitalization over 6 months. Results are presented as odds ratios (ORs) and 95% confidence intervals (CIs). An interaction term was added to the models to obtain the women-to-men ratios of ORs and 95% CIs, which were used to quantify how the ORs differed between the sexes.

**Results:** A total of 1210 patients (513 women, 697 men) were included in this analysis. They had a mean age of 73.7 (SD 8.9) years. The mean CFS score was 4.1 (SD 1.2). The prevalence of frailty was 40.7% across all participants, 46.8% in women vs. 36.3% in men, p<0.001. Women were older (mean age 74.6/SD 8.9 vs 73.0/SD 8.8 in men, p=0.004), had higher CHA=:JDS=:J-VASc score (4.2/SD 1.3 vs 3.2/SD 1.3 in men, p<0.001) and HASBLED score (1.6/SD 0.8 vs 1.4/SD 0.8 in men, p=0.004). OAC prescription rate was 90.7% across all participants and lowest among frail women (83.8%). With every unit increase in the CFS, the adjusted ORs for OAC prescription were 0.70 (95%CI 0.54–0.90) in women, 0.97 (95%CI 0.74–1.28) in men (women-to-men ratio of ORs 0.72, 95%CI 0.50–1.05). During the 6-month follow-up, the CVD hospitalization rate was 14.3%. CFS score predicted CVD hospitalization, with adjusted ORs of 1.65 (95%CI 1.29–2.10) in women, 2.04 (95%CI 1.62– 2.59) in men; women-to-men ratio of ORs 0.81 (95%CI 0.58–1.13).

**Conclusion:** In older patients with AF, frailty was more common and was associated with reduced odds of receiving OAC in women. Frailty increased the risk of CVD hospitalization for both women and men, although its impact on hospitalization tends to be greater in men. Further studies are needed to confirm these findings.

## Introduction

Atrial fibrillation (AF) is the most common type of arrhythmia globally and significantly increases the risk of ischemic stroke.^1,2^ Strokes associated with AF are usually more severe, and result in death or long-term disability.^1,2^ The financial burden of AF-related strokes on healthcare systems is considerably higher when compared to non-AF strokes.^3^ The prevalence and incidence of AF increase significantly as people get older, placing a significant strain on healthcare systems worldwide.^3-6^ According to a study in 2017, the global prevalence of AF was 0.51% (37,574 million cases), with the most significant recent rise observed in countries with a middle socio-demographic index.^7^ A 2023 study estimated that the prevalence of AF in the Asia-Pacific region was approximately 80 million individuals, significantly higher than the figures for other global regions.^8^

The management of AF includes stroke prevention through oral anticoagulants (OAC), symptom reduction with rate-control or rhythm-control strategies, and managing associated chronic health conditions.^1,3,6,9^ The use of OAC for stroke prophylaxis is essential in the management of AF, and studies have demonstrated the benefits of OAC in elderly populations.^2,3^ However, OAC has been reported to be underused in older people with AF, especially in those with frailty.^8,10,11^ Frailty has been shown to influence the prescriptions of OAC for stroke prevention, as well as patient outcomes.^12,13^ Frailty, defined as a clinical syndrome characterised by increased vulnerability to stressors and reduced physiological reserve, is very common among older adults, and it appears to occur more frequently, and with greater severity, in women compared to men.^14-17^ This difference may be partly attributed to women’s longer life expectancy, which results in a higher prevalence of age-related disorders.^18,19^ In addition, differences in muscle mass, hormonal profiles, and the burden of chronic disease can contribute to this disparity.^18-20^ Frailty is a strong predictor of poor outcomes in AF, including higher mortality, major bleeding events, hospitalization, and disability.^21,22^

Sex differences in frailty can significantly influence the treatment and outcomes of older adults with chronic health conditions. A male-female health-survival paradox has been described in the literature, which means that women, although having higher level of frailty, seem less vulnerable to death and adverse outcomes compared to men of the same age.^23^ In a meta-analysis including 37,426 participants from five studies in 2010, Gordon and colleagues reported that women exhibited a higher Frailty Index scores compared to men across all age groups.^14^ Despite this apparent disadvantage, the data from these studies revealed an interesting trend: females seem to manage their frailty more effectively than their male counterparts, as evident from their relatively lower mortality rates when compared to men with similar levels of frailty or age.^14^ The sex-frailty paradox has also been observed in several studies in older adults with heart failure. In a 2022 study of 499 older adults with heart failure (mean age 81 years) in Spain, although women had higher prevalence of frailty than men, frailty was found to be an independent predictor for 1-year mortality in men only.^24^ Another study published in 2023, which involved 115 older adults (mean age 64 years) with heart failure in the USA, found a higher prevalence of frailty in women, and frailty was identified as a significant predictor for the composite outcome of cardiovascular hospitalization and 1-year all-cause mortality for men but not women.^25^

There is limited evidence on sex differences in the impact of frailty on the treatment and outcomes in older adults with AF, although differences in clinical presentation, treatment, and outcomes of AF have been observed between women and men.^26,27^ Women with AF are often older at diagnosis than men and have more comorbidities, which can amplify frailty.^27^

Additionally, they are more likely to experience atypical AF symptoms, potentially delaying diagnosis or treatment.^27,28^ Some studies reported that frail women were more prone to ischemic strokes and also had higher risk of bleeding due to anticoagulants.^29,30^ Sex-specific data on frailty in older adults with AF is needed, and understanding sex differences is crucial for optimizing AF management in older adults.

Therefore, this study aimed to examine sex differences in the prevalence of frailty in older patients with AF and the association between frailty with OAC prescription and hospitalization, using observational study data from Vietnam, a middle-income country in Asia.

## Methods

### Study design and population

This study used pooled data from two prospective observational studies in Vietnam between December 2022 and May 2024. Older patients aged ≥ 60 diagnosed with AF who visited the clinics during the study period were recruited. AF was defined by medical history and confirmed with a 12-lead ECG. Patients with valvular heart disease or having an ischemic stroke within the past 2 weeks were excluded.

Data were collected from patient interviews and medical records. Information obtained included demographic characteristics, height, weight, medical history, and comorbidities. Body mass index (BMI) was calculated from measured weight and height, and classified into four goups: underweight (BMI < 18.5 kg/m^2^), ideal (BMI 18.5–22.9 kg/m^2^), overweight (BMI 23.0–24.9 kg/m^2^), and obese (BMI ≥ 25.0 kg/m^2^).^31^ Information of comorbidities including hypertension, dyslipidemia, diabetes, heart failure, chronic kidney disease, ischemic stroke, peripheral artery disease, hyperthyroidism, cancer, chronic obstructive pulmonary disease, chronic bronchitis were obtained from participants’ medical records.

CHA2DS2-VASc scores and HASBLED scores were calculated for all participants to assess the risk of ischemic stroke and bleeding, respectively.^1,2^ Frailty was defined by the Clinical Frailty Scale (CFS).^16,32^ The CFS score ranges from 1-9, and a cut-point of 5 was applied to define frailty.^32,33^

The studies were approved by the Ethics Committee of the University of Medicine and Pharmacy at Ho Chi Minh City (Reference Number 1027/HDDD-DHYD, dated 09/12/2022) and the Ethics Committee of Thong Nhat Hospital (Reference Number 87/2022/BVTN-HĐYĐ, dated 25/11/2022). Informed consent was obtained from all participants.

### Outcomes

The study outcome was the prescription of oral anticoagulants at discharge, and cardiovascular disease (CVD) related hospitalization over 6 months. Hospitalisation information was obtained from patient medical records and by making phone calls to the participants or their caregivers.

### Statistical analysis

Participant characteristics are presented as mean and standard deviation (SD) for continuous variables, and frequencies and percentages for categorical variables. Comparisons among groups (male/female, frail/non-frail) were conducted using chi-square tests or Fisher’s exact tests for binary variables, and Student’s t-tests or one-way ANOVA for continuous variables.

Logistic regression models were applied to examine the association between the CFS score with OAC prescription and CVD-related hospitalization over 6 months. Models with OAC as the outcome variable were adjusted for age, CHA=:JDS=:J-VASc score, and HASBLED score. Models with CVD-related hospitalization as the outcome were adjusted for age, CHA=:JDS=:J-VASc score, HASBLED score, and OAC prescription. Results are presented as odds ratios

(ORs) and 95% confidence intervals (CIs). An interaction term was added to the models to obtain the women-to-men ratios of ORs and 95% CIs, which were used to quantify how the ORs differed between the sexes.^34^ P values <0.05 were considered statistically significant. Data were analysed in SPSS Statistics 27.0.

## Results

A total of 1210 patients (513 women, 697 men) were included in this analysis. They had a mean age of 73.7 (SD 8.9) years. The mean CFS score was 4.1 (SD 1.2), higher in women (4.3, SD 1.3) compared to men (4.0, SD 1.2). Using a cut-point of 5, the prevalence of frailty was 40.7% overall, 46.8% in women vs. 36.3% in men, p < 0.001.

### Baseline data

Table 1 presents participant characteristics, stratified by sex. Women were older (mean age 74.6, SD 8.9 vs. 73.0, SD 8.8 in men, p = 0.004), had higher CHA=:JDS=:J-VASc score (4.2, SD 1.3 vs. 3.2, SD 1.3 in men, p < 0.001) and HASBLED score (1.6, SD 0.8 vs. 1.4, SD 0.8 in men, p=0.004). The prevalence of underweight was significantly higher in women (11.7%) compared to men (8.5%). Among comorbidities, women had higher prevalence of heart failure (29.4% compared to 22.5% in men), hyperthyroid (3.9% compared to 1.4% in men), while men had higher prevalence of hypertension (96.6% vs. 94.2% in women), cancer (2.9% vs. 0.6% in women), and chronic pulmonary disease (2.9% vs. 0.6% in women).

**Table 1.**
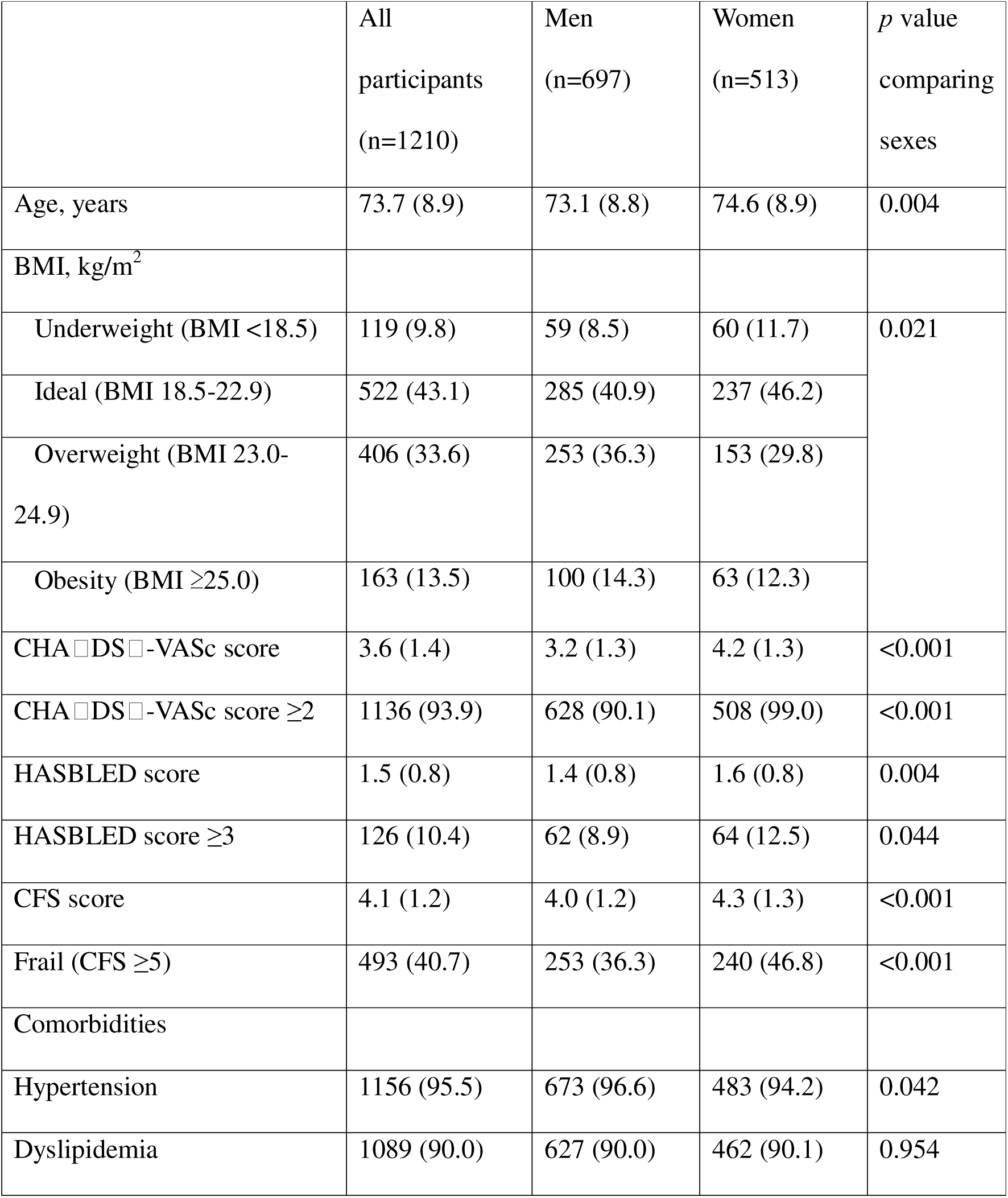
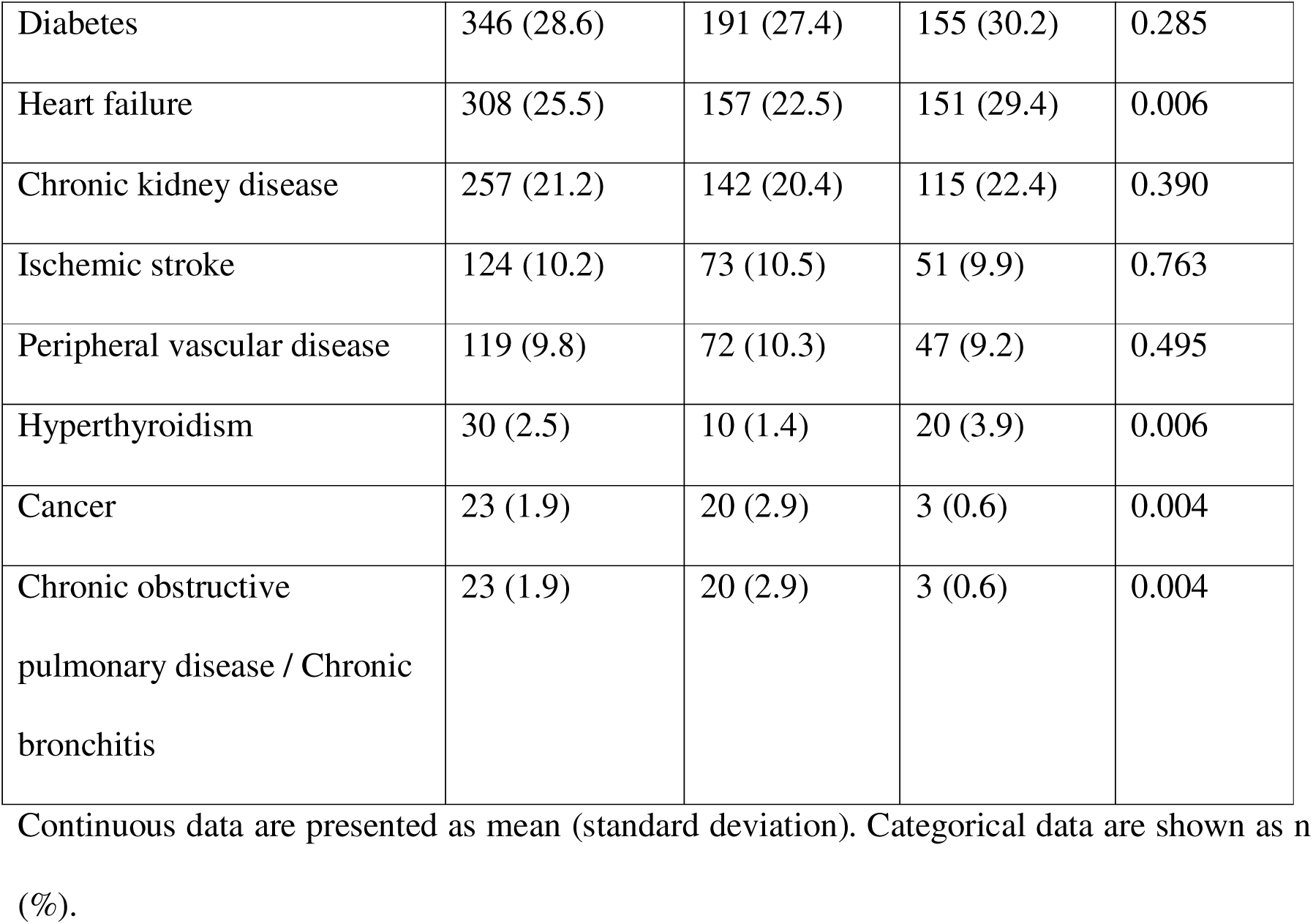
Participant characteristics.

|  | All<br>participants<br>(n=1210) | Men<br>(n=697) | Women<br>(n=513) | *p* value<br>comparing<br>sexes |
| --- | --- | --- | --- | --- |
| Age, years | 73.7 (8.9) | 73.1 (8.8) | 74.6 (8.9) | 0.004 |
| BMI, kg/m<sup>2</sup> |  |  |  |  |
| Underweight (BMI <18.5) | 119 (9.8) | 59 (8.5) | 60 (11.7) | 0.021 |
| Ideal (BMI 18.5-22.9) | 522 (43.1) | 285 (40.9) | 237 (46.2) |  |
| Overweight (BMI 23.0-24.9) | 406 (33.6) | 253 (36.3) | 153 (29.8) |  |
| Obesity (BMI ≥25.0) | 163 (13.5) | 100 (14.3) | 63 (12.3) |  |
| CHA<sub>2</sub>DS<sub>2</sub>-VASc score | 3.6 (1.4) | 3.2 (1.3) | 4.2 (1.3) | <0.001 |
| CHA<sub>2</sub>DS<sub>2</sub>-VASc score ≥2 | 1136 (93.9) | 628 (90.1) | 508 (99.0) | <0.001 |
| HASBLED score | 1.5 (0.8) | 1.4 (0.8) | 1.6 (0.8) | 0.004 |
| HASBLED score ≥3 | 126 (10.4) | 62 (8.9) | 64 (12.5) | 0.044 |
| CFS score | 4.1 (1.2) | 4.0 (1.2) | 4.3 (1.3) | <0.001 |
| Frail (CFS ≥5) | 493 (40.7) | 253 (36.3) | 240 (46.8) | <0.001 |
| Comorbidities |  |  |  |  |
| Hypertension | 1156 (95.5) | 673 (96.6) | 483 (94.2) | 0.042 |
| Dyslipidemia | 1089 (90.0) | 627 (90.0) | 462 (90.1) | 0.954 |
| Diabetes | 346 (28.6) | 191 (27.4) | 155 (30.2) | 0.285 |
| Heart failure | 308 (25.5) | 157 (22.5) | 151 (29.4) | 0.006 |
| Chronic kidney disease | 257 (21.2) | 142 (20.4) | 115 (22.4) | 0.390 |
| Ischemic stroke | 124 (10.2) | 73 (10.5) | 51 (9.9) | 0.763 |
| Peripheral vascular disease | 119 (9.8) | 72 (10.3) | 47 (9.2) | 0.495 |
| Hyperthyroidism | 30 (2.5) | 10 (1.4) | 20 (3.9) | 0.006 |
| Cancer | 23 (1.9) | 20 (2.9) | 3 (0.6) | 0.004 |
| Chronic obstructive<br>pulmonary disease / Chronic<br>bronchitis | 23 (1.9) | 20 (2.9) | 3 (0.6) | 0.004 |

Continuous data are presented as mean (standard deviation). Categorical data are shown as n (%).

Figure 1 illustrates the mean CHA=:JDS=:J-VASc score and HASBLED score across the four groups of frail women, frail men, non-frail women, and non-frail men. Frail women had the highest risk of stroke (mean CHA=:JDS=:J-VASc score 5.0, SD 1.2) and bleeding (mean HASBLED score 1.8, SD 0.8), followed by frail men (mean CHA=:JDS=:J-VASc score 3.9, SD 1.2; mean HASBLED score 1.7, SD 0.8), non-frail women (mean CHA=:JDS=:J-VASc score 3.6, SD 1.1; mean HASBLED score 1.4, SD 0.8), and non-frail men (mean CHA=:JDS=:J-VASc score 2.8, SD 1.2; mean HASBLED score 1.3, SD 0.7)

**Figure 1.**
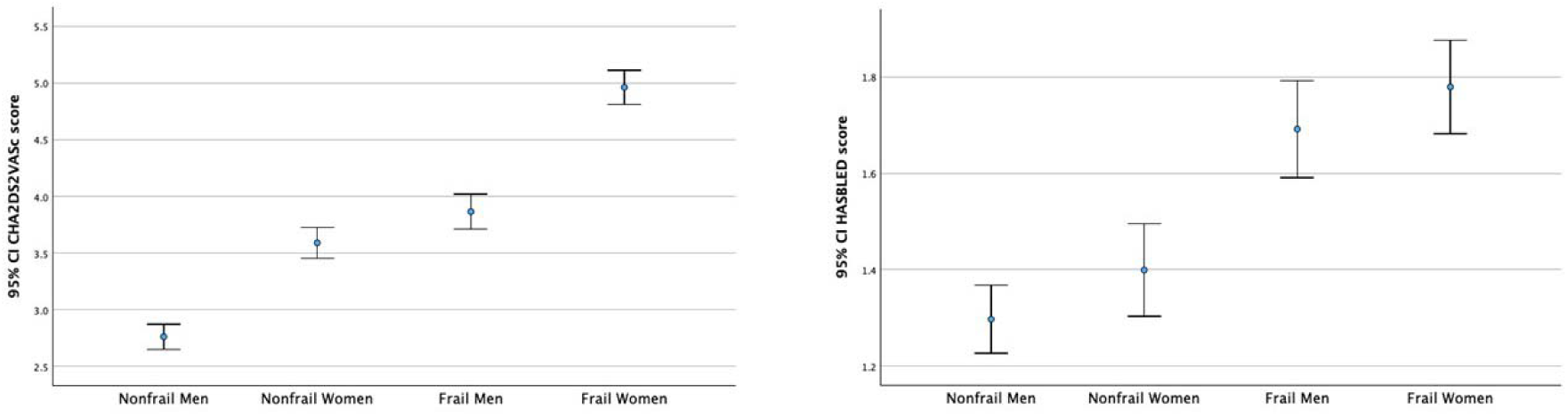
Mean CHADDSD-VASc score and HASBLED score by sex and frailty.

### Anticoagulant prescription at discharge

At discharge, the OAC prescription rate was 90.7% in all participants, 87.9% in women vs. 92.7% in men (p=0.005). The prescription rate of OAC was lower in women vs. men in the frail group (83.8% vs. 92.5%, p = 0.003). There was no significant difference between women and men in the non-frail group (Figure 2). After adjustments, for every unit increase in the CFS the ORs for OAC prescription were 0.70 (95% CI 0.54 – 0.90) in women, 0.97 (95% CI 0.74 – 1.28) in men (women-to-men ratio of ORs 0.72, 95% CI 0.50 - 1.05). (Table 2)

**Figure 2.**
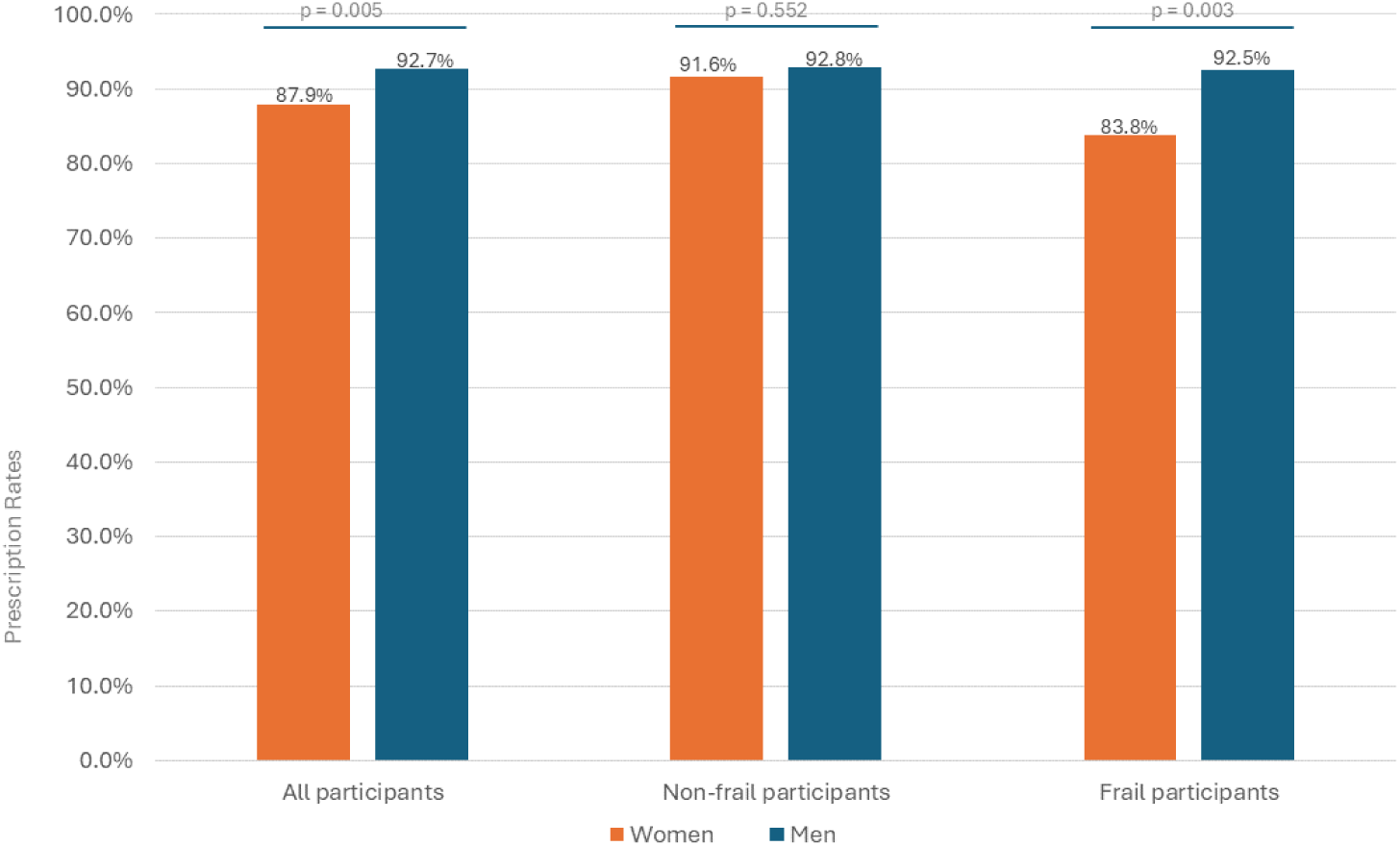
Anticoagulant prescription rates by sex and frailty.

**Table 2.**
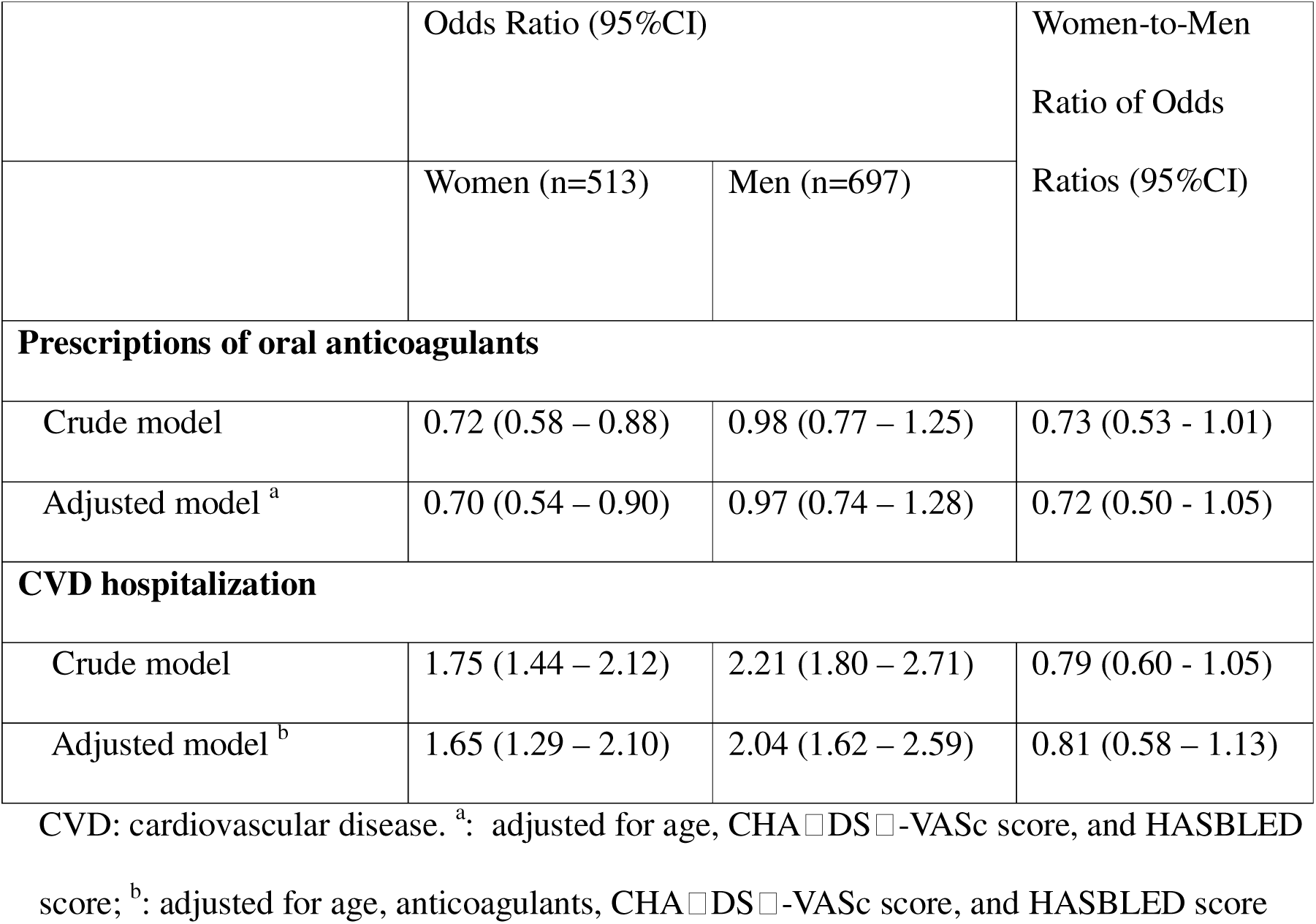
Sex differences in the association of the Clinical Frailty Scale with anticoagulant prescriptions and 6-month cardiovascular-related hospitalization.

|  | Odds Ratio (95%CI) |  | Women-to-Men<br>Ratio of Odds<br>Ratios (95%CI) |
| --- | --- | --- | --- |
|  | Women (n=513) | Men (n=697) |  |
| **Prescriptions of oral anticoagulants** |  |  |  |
| Crude model | 0.72 (0.58 – 0.88) | 0.98 (0.77 – 1.25) | 0.73 (0.53 - 1.01) |
| Adjusted model <sup>a</sup> | 0.70 (0.54 – 0.90) | 0.97 (0.74 – 1.28) | 0.72 (0.50 - 1.05) |
| **CVD hospitalization** |  |  |  |
| Crude model | 1.75 (1.44 – 2.12) | 2.21 (1.80 – 2.71) | 0.79 (0.60 - 1.05) |
| Adjusted model <sup>b</sup> | 1.65 (1.29 – 2.10) | 2.04 (1.62 – 2.59) | 0.81 (0.58 – 1.13) |

CVD: cardiovascular disease. <sup>a</sup>: adjusted for age, CHA<sub>2</sub>DS<sub>2</sub>-VASc score, and HASBLED score; <sup>b</sup>: adjusted for age, anticoagulants, CHA<sub>2</sub>DS<sub>2</sub>-VASc score, and HASBLED score

### Cardiovascular hospitalization at 6-month follow-up

During the 6-month follow-up, the CVD hospitalization rate was 14.3% in all participants, 17.0% in women vs. 12.3% in men (p=0.022), with no significant differences among women and men in the frail and non-frail sub-groups (Figure 3). In the regression models, with every unit increase in the CFS score, the adjusted ORs for CVD hospitalization were 1.65 (95% CI 1.29 – 2.10) in women, 2.04 (95% CI 1.62 – 2.59) in men, women-to-men ratio of ORs 0.81 (95% CI 0.58 – 1.13). (Table 2)

**Figure 3.**
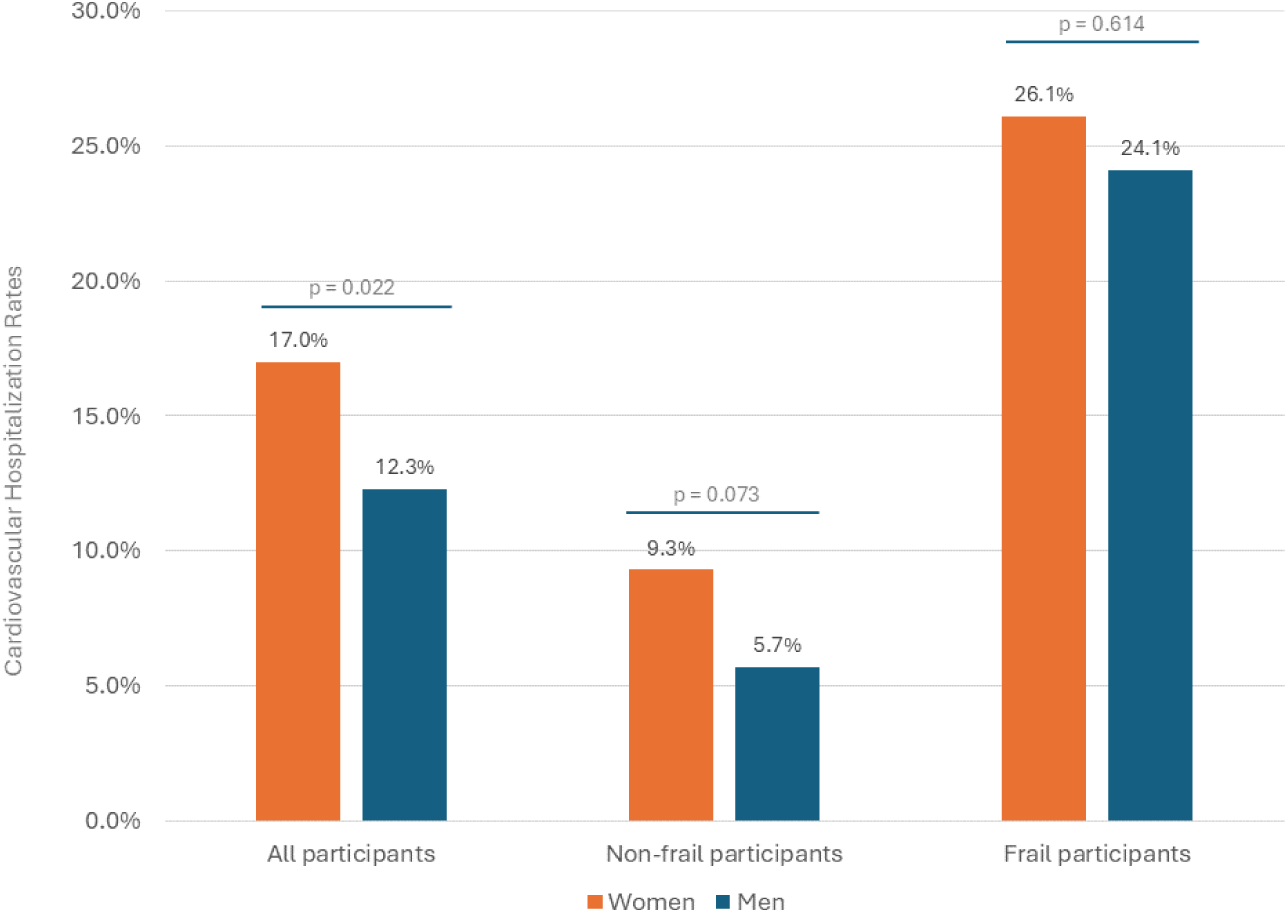
Cardiovascular hospitalization rates by sex and frailty.

## Discussion

In 1210 Vietnamese participants aged 60 years or older with AF, we found a high prevalence of frailty. Compared to men, women had a higher burden of frailty, stroke risk and bleeding risk. Frailty reduced the odds of receiving OAC in women, but not in men. Frailty increased the risk of CVD hospitalization for both women and men, although its impact on hospitalization tended to be greater in men.

The reasons why frailty was associated with reduced odds of anticoagulant prescription in women with AF compared to men can be multifaceted, involving socio-economic, physiological, clinical, and systemic factors. Women with frailty are usually older and have lower body weight, reduced renal function, and polypharmacy compared to men.^14,20^ Frailty can increase the risk of adverse effects of anticoagulant, as it is associated with reduced physiological reserve and impaired pharmacokinetics and pharmacodynamics.^14,20^ Frailty also increases the risk of falls, a major concern in anticoagulant therapy due to potential intracranial or major bleeding.^35^ Compared to their male counterparts, older women are at higher risk of falls due to lower muscle mass, balance disorders, and osteoporosis.^36,37^ Therefore, physicians may perceive frail women as having a higher risk of adverse events from anticoagulant (such as major bleeding and falls) compared to frail men, leading to greater caution in prescribing OACs for them. In fact, a meta-analysis involving 1,085,931=:Jwomen and 1,387,123 men in 2024 showed that women may have higher gastrointestinal bleeding risk with direct oral anticoagulants compared to men.^26^ Further studies are needed to understand prescribers’ perspectives on OAC prescription for stroke prevention in frail older women with AF. Frailty is a critical yet often overlooked geriatric condition in post-discharge care for older patients, particularly for women. Clinicians should prioritize frailty assessments and incorporate frailty into post-discharge care plans. Post-discharge care must also address the social and psychological dimensions of frailty. Compared to men, women may face unique socioeconomic challenges, particularly for women living in low- and middle-income countries.^38-40^

The finding that frailty increased the risk of CVD hospitalization in our study is consistent with previous studies worldwide.^41,42^ In our study, although women were frailer and at higher risks of both strokes and bleeding, the impact of frailty on CVD hospitalization over 6 months tended to be stronger in men. Although the sex difference in the impact of frailty on CVD hospitalization observed in our study were not statistically significant, they are highly suggestive of important variations that deserve further investigation. More research is needed to understand the mechanisms for this difference. Our findings may support the observed sex-frailty paradox in older adults. While women appear to be frailer, they demonstrate a remarkable resilience that translates into longer survival rates. This paradox may suggest a complex interaction between biological, socio-economic, and possibly behavioural factors that contribute to the differing outcomes observed between men and women.^23,43^

As data on sex-frailty interactions in older adults with AF remain limited, our findings reinforce the concept of sex differences in frailty related to AF in particularly and health in general, and highlight the need for further investigation into the underlying mechanisms that contribute to this paradox. Frailty can amplify the challenges of managing AF in older adults, with women often bearing a heavier burden due to higher frailty prevalence and distinct risk profiles. Effective treatment requires a personalized approach that carefully balance the risks of bleeding and stroke. To achieve the best outcomes, it is crucial to address frailty alongside AF-specific therapies. Understanding these factors could enhance the effectiveness of health interventions and strategies aimed at reducing adverse events for men, while improving overall health outcomes for both sexes. Frailty assessment should be conducted routinely for older adults with AF, and women might benefit from earlier frailty screening given their higher baseline risk.

To the best of our knowledge, this is the first study to explore sex differences in the impact of frailty on OAC prescription and CVD-related hospitalization in an older Asian population with AF. Our study provides data specific to Vietnamese patients with AF, where research on frailty in this population remains limited. However, this study is without limitations. The study was conducted at the outpatient clinics of only two urban hospitals. The follow up period was short, spanning just six months, which may limit our insights on the impact of frailty on the risk of hospitalizations in women and men. Therefore, the findings may not be generalisable for all older adults with AF in Vietnam and should be interpreted with caution.

## Conclusion

In older patients with AF, frailty was more common and was associated with reduced odds of receiving anticoagulants in women. Frailty increased the risk of CVD hospitalization for both women and men, although its impact on hospitalization tends to be greater in men. These findings align with the observed sex-frailty paradox in older adults in general. Further research is needed to confirm these findings and to understand the mechanisms underlying this discrepancy.

## Data Availability

All data produced in the present study are available upon reasonable request to the authors.

